# SARS-CoV-2 receptor mutation in Egyptian population

**DOI:** 10.1101/2020.04.27.20082206

**Authors:** Mohamed M Hafez, Zeinab K Hassan, Abeer A. Bahnasy, Ola S. Ahmed, Mohamed Abouelhoda, Abdel-Rahman N. Zekri

## Abstract

The Coronavirus disease 2019 (COVID-19) is a respiratory tract infectious disease caused by Severe Acute Respiratory Syndrome coronavirus 2 (SARS-CoV-2). SARS-CoV-2 triggers severe pneumonia leading to acute respiratory distress syndrome and death in severe cases. According to WHO reported, Egypt is among the countries with low confirmed SARS CoV2 infected symptomatic cases and death. We postulate that one of the reasons for this may be due mutations in the viral receptor. Therefore this study was conducted to confirm or reject this postulation.

## Manuscript

The Coronavirus disease 2019 (COVID-19) is a respiratory tract infectious disease caused by Severe Acute Respiratory Syndrome coronavirus 2 (SARS-CoV-2). SARS-CoV-2 triggers severe pneumonia leading to acute respiratory distress syndrome and death in severe cases. According to WHO reported, Egypt is among the countries with low confirmed SARS CoV2 infected symptomatic cases and death. We postulate that one of the reasons for this may be due mutations in the viral receptor. Therefore this study was conducted to confirm or reject this postulation. Genomic DNA was extracted from 70 peripheral blood samples from Egyptian individual’s collected long time before the pandamic. The whole exome sequencing was done using next generation sequencing as sequences of 100 bp paired-end reads library on Ion Torrent platform. Then, the data were filtered out to remove the low-quality reads forming clean data in which bioinformatics analyses were done. The ACE2 and BACE2 Alp 56 sequences were analyzed using bioinformatics software to detect the difference mutations among Egyptian population in comparison to international population databases. Our data concluded that no SNPs or mutations were detected in ACE2 or BACE2 Alp 56 receptor among Egyptian population, hence no correlation with the low symptomatic incidence/death of SARS-Cov-2 viruses in the Egyptian populations.

The SARS-CoV-2 causes a new type of pneumonia that forms a global public health problem. As of April 21, 2020, more than 2,300,000 confirmed COVID-19 cases were reported worldwide with 162 956 confirmed deaths. Angiotensin-converting enzyme 2 (ACE2) and Beta-site amyloid beta A4 precursor protein-cleaving enzyme 2 (BACE2 Alp 56) have been identified as the main receptor for SARS-CoV-2. ACE2 is normally expressed in cardiovascular, lung type II alveolar epithelial cells and kidneys with tissue-specific activity patterns [1, 2] and also in the placenta [3]. The serine protease for virus Spike (S) protein priming, TMPRSS2, was also reported to be necessary for SARS-CoV-2 cell entry [4]. ACE2 is a Renin-angiotensin system (RAS) component and controls blood pressure [5]. The angiotensin-converting enzyme-2, which is encoded by *ACE2* gene, is the receptor for SARS-CoV-2 [7, 8]. Some reports showed a positive correlation between SARS-CoV infection level and ACE2 expression levels *in vitro* [9, 10]. The susceptibility to infection with SARS-CoV-2 may depend on ACE2 expression level and pattern in different tissues and also may be among different populations, in which the *ACE2* gene sequence or expression levels need further studies.

This experiment was performed to investigate the co-relationship between the low incidence of SARS-CoV-2 and ACE2 receptor among Egyptian population. The current study was done in accordance with The Code of Ethics of the World Medical Association of Helsinki Declaration [6] and was approved by the Ethics Committee of National Cancer Institute, Cairo University. All participants signed a written informed consent. Genomic DNA was extracted from 70 peripheral blood samples from normal Egyptian participants using a QIAamp DNA Mini kit (Qiagen GmbH) according to manufacturer’s instructors. In order to construct a library, exome capture was performed using an Agilent SureSelect Human All ExonV5 kit (Agilent Technologies, Inc.) according to manufacturer’s protocol. In brief, a total of 0.5 µg DNA was used for DNA library preparations followed by fragmentation via sonication to ~350 bp size. Then, DNA fragments were end polished, A-tailed and ligated with full-length adapter for sequencing, followed by polymerase chain reaction (PCR) for amplification of libraries with full-length adapters using KAPA HiFi HotStart ReadyMix (Roche Diagnostics) with P5: 5′-AATGATACGGCGACCACCGAGATC-3′ and P7: 5′-CAAGCAGAAGACGGCATACGA-3′ primers according to standard protocol. The qualified library was sequenced as 100 bp paired-end reads on Ion Torrent platform (Applied Biosystems; Thermo Fisher Scientific, Inc.) according to the manufacturer's protocols.

We found one frameshift mutation in exon 15 (c.1995 delT), one synonymous NSP in exon16 (c.2070 T>C) and three splicing in exon18 (NM_021804 TA>AG, NM_021804 CT>GG and NM_021804 CT>GG) and NM_021804:exon4:c.439+4G>A in ACE2 receptor. On the other hand, we found 9 SNPs in BACE2 receptor, which are 3 non synonymous SNP in exons 1, 3 and 8 (c.190 C>G, c.455 G>A and c.1156 A>T, respectively) 2 synonymous SNP in exons 1 and 7 (c.192 G>C and c.1092 C>T), one frameshift mutation in exon1 (c.216 delG) and exon4:c.619–5C>T, exon5:c.748–14->T and exon5:c.748–10C>T. All the SNP variants are common in world population and their structural damaging scores are not high enough to affect the protein structure. Each of the two frameshift deletions has seen only once in our population and their quality is not strong enough to support structural changes in the protein (All data are available as supplement file).

We also compared our findings to the reported variants of the Italian population. The most relevant variants in the Italian population were p.Asn720Asp, p.Lys26Arg, p.Gly211Arg, and p.Trp69Cys. The first three variants are rare in world population in general (ExAC All 0.016, 0.001, 0.005, respectively) but the frequency in African, Middle East, and East Asian populations is much less than that of European population. Their structural damaging scores are not above the accepted threshold (CADD less than 15, and Benign PolyPhen and Sift predictions). The fourth variant is the most important and it is specific to the Italian population with high structural damaging score (CADD =23). All these four variants do not exist in our exome set of the Egyptian population.

In conclusion and as per available data, mutations in the SARS-CoV-2 receptor were not the reason for the low symptomatic incidence/death rate of the Covid-19 in Egypt. We recommend a large sample size study on the host and viral genome sequences, including potential functional coding variants in *ACE2* and the gene expression levels, among Egyptian populations for further epidemiological investigations of SARS-CoV-2.

## Data Availability

Supplementary file attached.

## Notes

### Competing Interest Statement

The authors have declared no competing interest.

### Funding Statement

work was supported by STDF grant.

